# Internet-based tool for visualizing county and state level COVID-19 trends in the United States

**DOI:** 10.1101/2020.05.11.20095851

**Authors:** S Hinz, MM Basam, KY Aguilera, MA LaBarge

**Affiliations:** Department of Population Sciences, Beckman Research Institute, City of Hope, 1500 E. Duarte Rd, Duarte, CA 91010, USA; Department of Pathology and Laboratory Medicine, David Geffen School of Medicine at UCLA, Los Angeles, CA, 90095, USA

## Abstract

The novel COVID-19 outbreak started in 2019 in Wuhan China and quickly spread to at least 185 countries. We developed an interactive web application that allows users to visualize the spread of COVID-19 in the Unites States at state and county levels. This tool allows visualization of how the virus spreads over time and how state-wide efforts to reduce transmissions have affected the curve in local areas. The downloadable application data allows users to conduct additional analyses. We demonstrate exemplars of trend analyses comparing the daily infection and death rates before and after safer at home orders were implemented per state. The goal was to develop a COVID-19 tracking tool that informs users about the spread of the virus to enable them to make informed decisions after better understanding the presented data.

## Introduction

In Wuhan (Hubai, China) a cluster of pneumonia infections occurred in December of 2019. In January 2020 a novel coronavirus SARS-CoV-2 was identified as the cause in patient samples from Wuhan (Zhu et al., 2020). This outbreak has since spread to at least 185 countries and infected over 3 million people (as of May 1, 2020). Response to the pandemic in the United States is coordinated mainly at the state government level. In order to track and visualize the spread of coronavirus disease (COVID-19) and evaluate trends in response to state government interventions in the United States, we developed an automatically updating Shiny application. This application enables users to view trends in numbers of verified COVID-19 infections and deaths as reported by county health departments across the United States.

## Methods

### Data

The county-level and state-level data for COVID-19 infections and deaths is obtained from *The New York Times (NYT)* ongoing GitHub repository (https://github.com/nytimes/covid-19-data). Their data are compiled from state and local government and health department reports and is updated daily around 17:00 PST. State government non-pharmaceutical intervention data were provided through Killeen et al. (2020). County government non-pharmaceutical intervention data were provided by The COVID Tracking Project and is accessible on their website (https://covidtracking.com/api). State and county population and demographic information is accessed through the U.S. Census Bureau. We use their 2018 population estimates to get a more accurate sense of current population levels and demographic breakdown.

### App Interface and Data visualization

We implemented the U.S. COVID Tracker app as an independently updating app on a shinyapps.io server that pulls the latest data from the NYT data repository. Once a user opens the app, it loads a map of the U.S. with daily updating counters on the left of the map, and tables and plots at the bottom of the map (Figure 1). The interactive map allows users to zoom, pan, and click on individual locations that can change based on their map view selection. By default, state-level data are shown with a pseudo-coloring according to the total number of confirmed infections in a respective state. By selecting county view, the map updates to show all counties in the U.S. with confirmed infections. The “normalize by population” option allows users to rescale the map’s color by normalized confirmed infections (per 100K residents). When a state or county is selected, a pop-up displays the number of confirmed infections, deaths, and demographic information about that locality. To the left of the map, we display updating counters that allow for a quick view of the cumulative number of confirmed infections and deaths, as well as a daily counter of both. Lastly, we display the date when the NYT data have been last updated.

**Figure 1:**
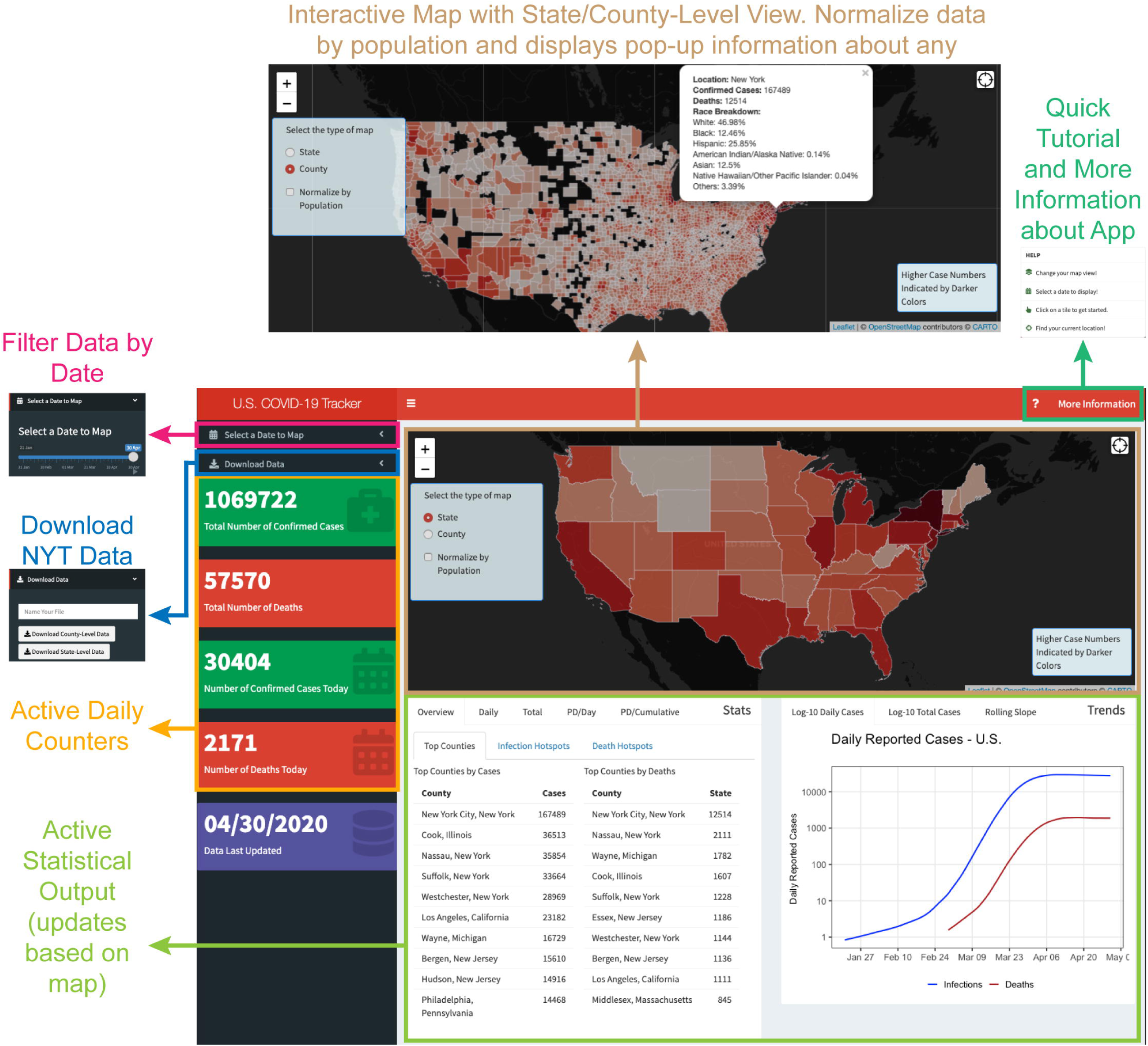
US COVID-19 Tracker interface. Overview of the COVID-19 tracker interface with magnification of the interactive map for state and county level infection data.

Below the map, we display multiple table and plot options for the user to view on a country, state, or county specific level to help with data interpretation. Upon opening the app, country-wide plots show the overall impact of COVID-19 on the U.S. (Figure 2). Upon interaction with the map, the plots are updated automatically to show state or county specific data. Additionally, we allow for users to filter the data based on previous dates, as well as normalize the number of confirmed infections or deaths by a state or county’s population (per 100K residents). We display two tables that show the top 10 counties in terms of their number of confirmed infections and deaths (Figure 2A) to show the most impacted counties. Next, we display the top 10 county hotspots based on their confirmed infections or death population doublings (Figure 2B-C). We filter for counties that have 10 or more confirmed infections or deaths and calculate population doublings as the latest count divided by the previous day’s count. The change between the previous day and latest day’s data are shown as well.

We display quick statistics on the left panel to allow users to quickly digest how COVID-19 has impacted the U.S., or a state or county of interest. We display the number of daily confirmed infections and deaths as well as their cumulative numbers with trendlines and bars where appropriate (Figure 2D-E). Additionally, we display log10 transformed daily and cumulative counts in the Trends subpanel to show the overall trend of the data (Figure 2G-H). Daily and cumulative population doublings are displayed in the Stats panel and are calculated by the following formulas: PD_daily_=log_10_(Day_i_/Day_i-1_)/log_10_(2) and PD_cumulative_=log_10_(Day_Σi_/Day_Σi-1_)/log_10_(2); respectively (Figure 2F & I).

**Figure 2:**
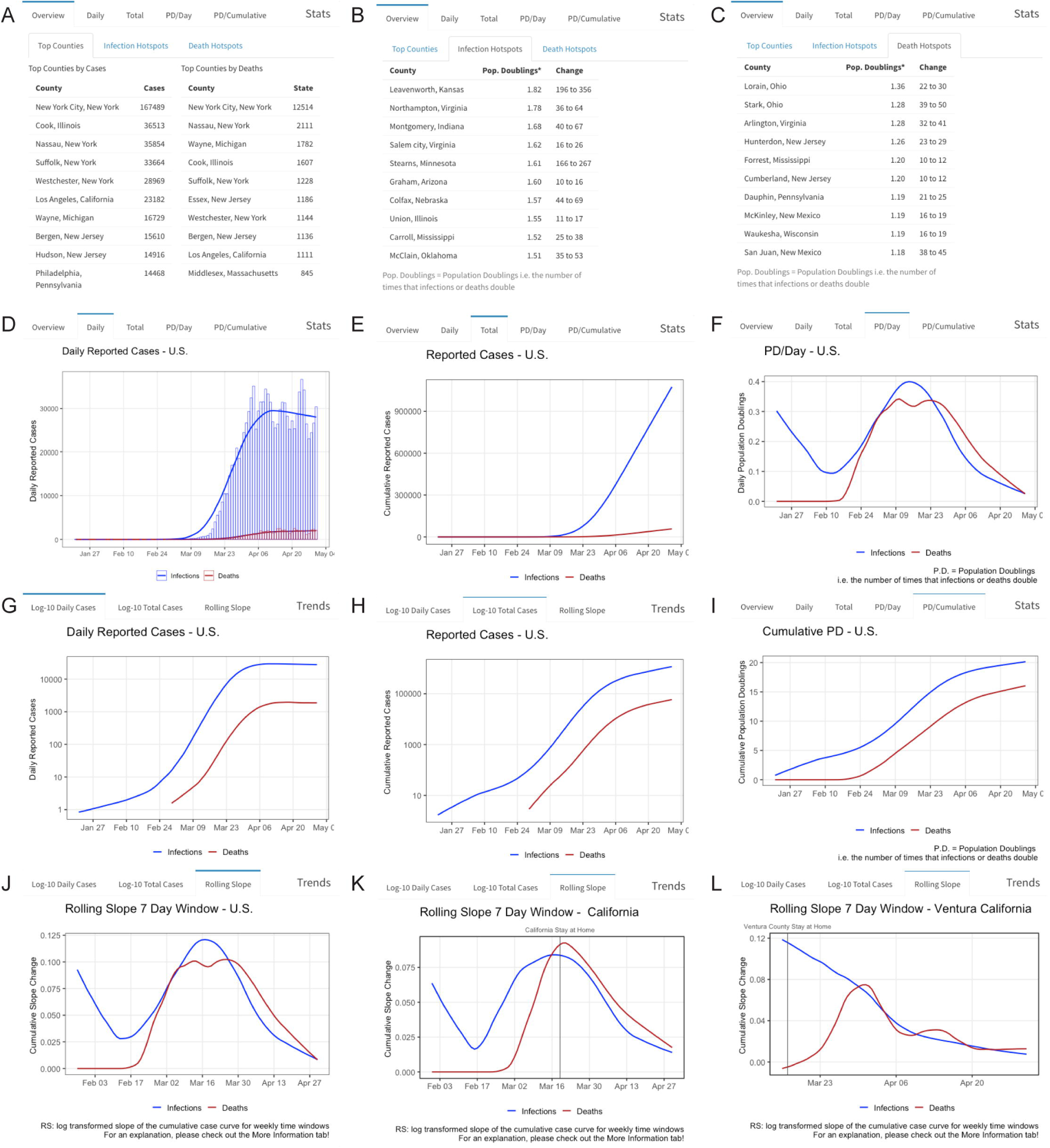
Example graph panels accessible through the COVID-19 application. A-C) Tables highlighting counties with highest numbers and hotspots for infections and deaths. Graphs depicting daily reported cases (D), cumulative reported cases (E), daily population doublings (F), and respective log10 transformations for the US and all counties (G-I). Rolling slope of the log10 transformed cumulative cases for reported infections and deaths (J-L).

In the Trends subpanel a rolling slope plot is displayed that allows for illustration of the “flattening of the curve”, or the slope changes of the log transformed cumulative death and infection numbers (Figure 2J-L). A moving 7-day window was applied to extract the slope of the linear regression as a function of 1-week intervals. These windows were calculated for every 7-day period and the “rolling slope” values were plotted against the date. Higher slopes are indicative of a surge in the number of confirmed infections or deaths whereas a slope of 0 indicates stagnant confirmed case or death numbers.

Additionally, we provide our users the ability to download the data, which includes the NYT COVID-19 data, as well as population statistics on each county or state (Figure 1). Users can view a quick tutorial on how to use the app by clicking on the question mark and the “More Information” tab allows them to read an in-depth explanation about the app, data, and functions used.

### Tool implementation

The program was written in R (R. Core Team, 2019) using R Studio, Shiny (Chang et al., 2020), shinydashboard (Chang and Ribeiro, 2018), leaflet (Cheng et al., 2019), tidyr (Wickham and Henry, 2020), ggplot (Wickham, 2016), dplyr (Wickham et al., 2020), zoo (Zeileis and Grothendieck, 2005), data.table (Dowle and Srinivasan, 2019), readr (Wickham et al., 2018), rgdal (Bivand et al., 2019), rintrojs (Ganz, 2016), janitor (Firke, 2020), and stringr (Wickham, 2019) packages. It is hosted through the authors’ private account in shinyapps.io.

## Results

### Applicability of the underlying data of the internet-based tool to assess trends

As a means to reduce the spread of COVID-19, national guidelines were set at both the county and state level. The implementation of safer at home orders, per state, resulted in effective flattening or decreasing of the rate of new infections and deaths per 100,000 residents in most states (Figure 3). 43 U.S. states set safer at home parameters based on local and state guidelines, whereas 7 U.S. states as of May 1, 2020 did not (Kansas, Iowa, Nebraska, North Dakota, Oklahoma, South Dakota, and Utah). A surrogate safer at home date was extrapolated for these 7 states from the average of national safer at home dates (March 27, 2020). The states that did not exercise safer at home orders have demonstrated variable changes to the confirmed new infections and deaths (Figure 4). However, an increase in new infections was demonstrated, especially in Arkansas, Iowa, Nebraska, North Dakota, and Utah. Comparisons of the rate of new infections of all 50 states, calculated as the difference in the slope from before and after the safer at home orders were implemented, show a decrease in newly confirmed infections (Figure 5). These data transformations are not presently incorporated in the app, but highlight the applicability of the underlying downloadable data.

**Figure 3:**
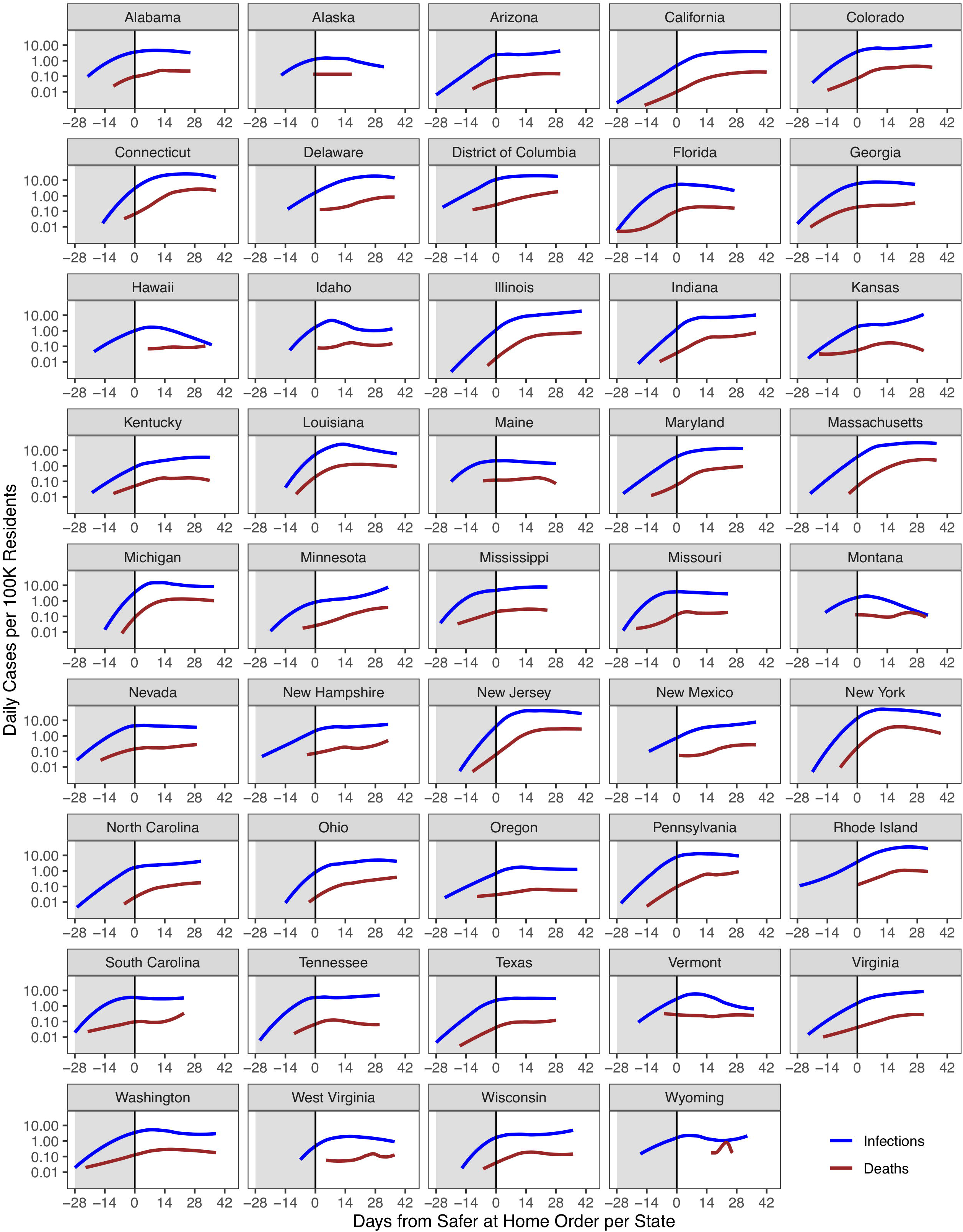
Application of internet-based tool’s data. Reported numbers of infections (blue) and deaths (red) per 100,000 residents per state plotted as a function of the days from the states’ safer at home order. Shaded area indicates 28-day period prior to the order.

**Figure 4:**
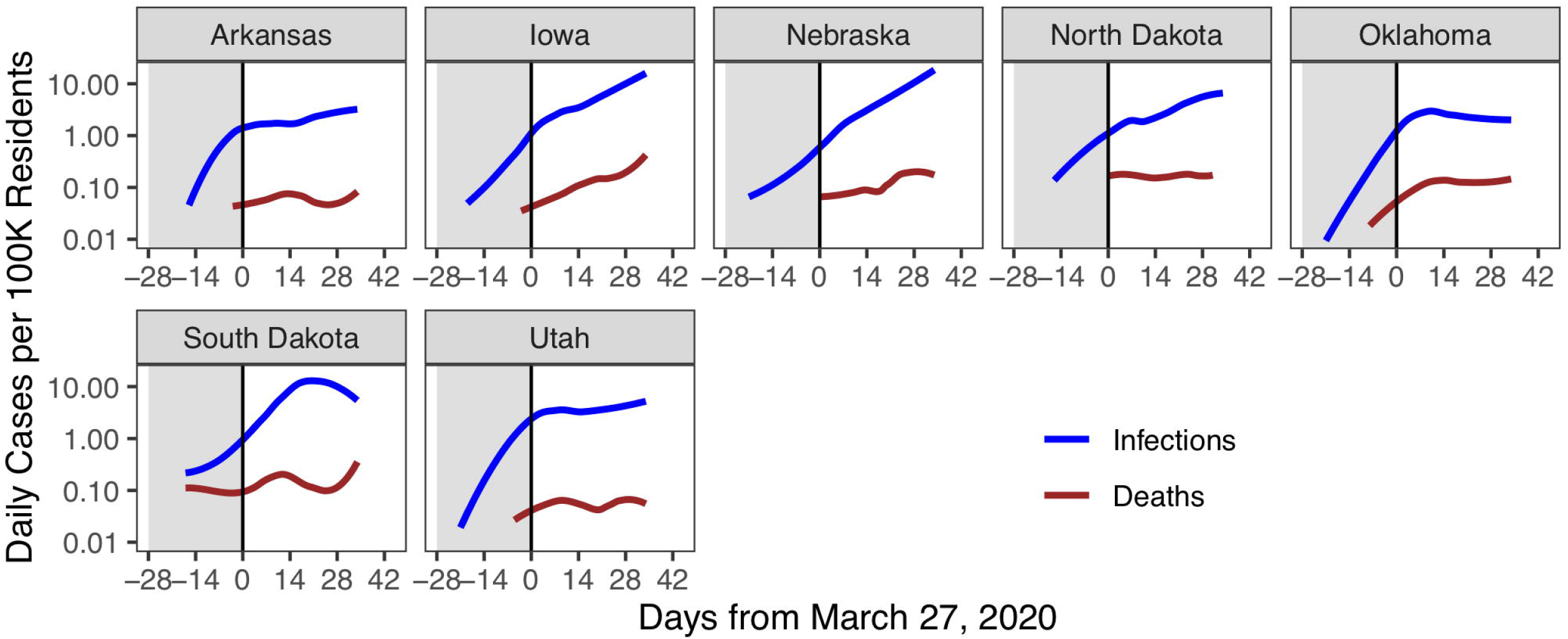
Application of internet-based tool’s data. Reported numbers of infections (blue) and deaths (red) per 100, 000 residents per state without a safer at home order. Cases are plotted as a function of the days from March 27, 2020 (national average of all state implementation dates). Shaded area indicates 28-day period prior to the date.

**Figure 5:**
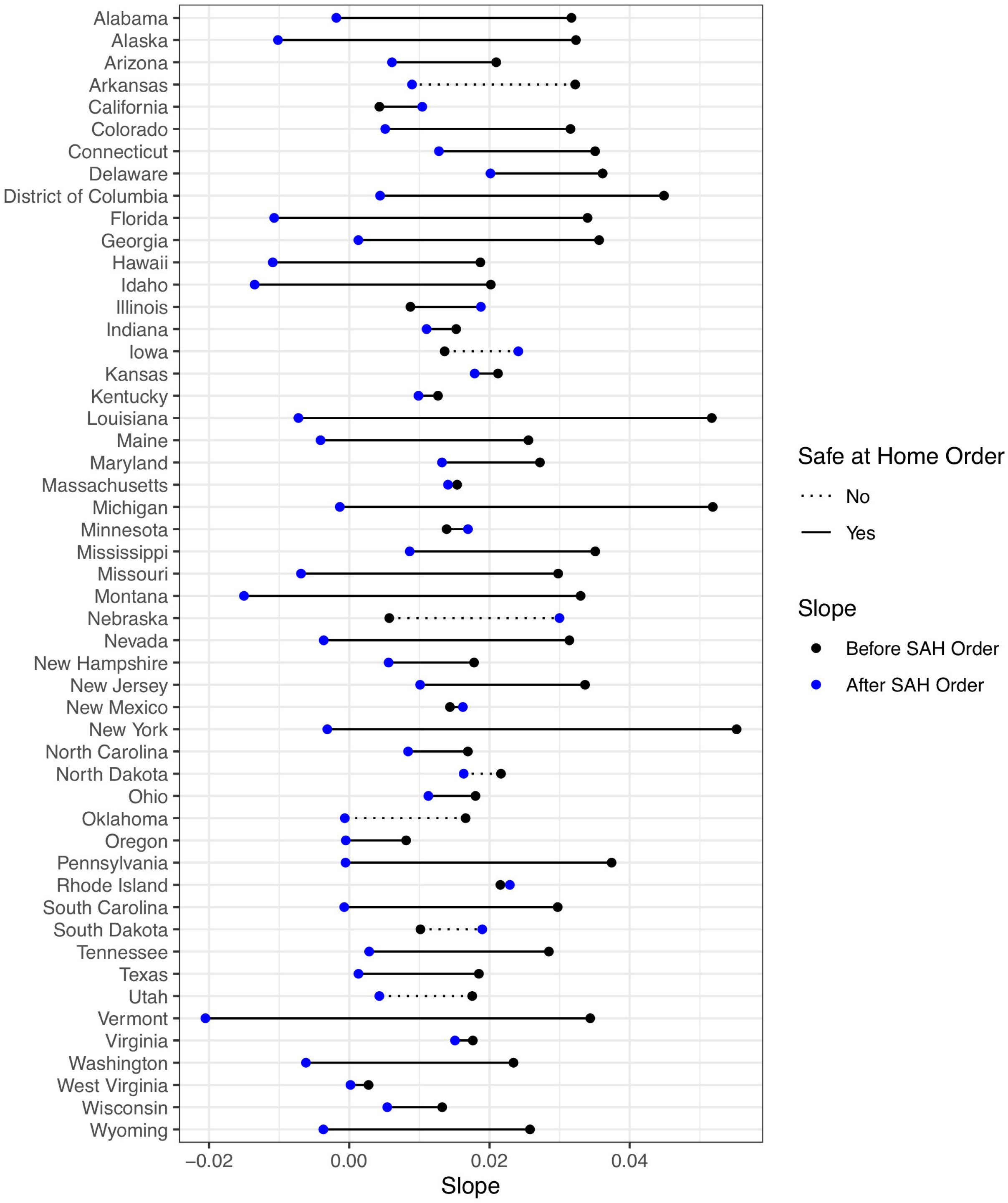
Assessment of trend changes based on the application’s downloadable data. Slope of the linear regression of the log10 transformed confirmed daily infections before (black) and after (blue) safer at home measures. For states without a safe at home order (dashed line) March 27, 2020 (national average of all state implementation dates) was used as the surrogate date.

## Discussion

The tool reports confirmed COVID-19 infections and deaths at the state and county level and is automatically updated daily. The interactive map of the US allows users to visualize the distribution of confirmed COVID-19 infections (Figure 1) with plots for summary statistics, data transformations, and trends for the US. Additionally, we implemented data visualization at the state and county level (Figure 1B). Upon user interaction with the map, the graphs update to reflect the data in the selected state or county (Figure 2 A-D), including annotations for statewide stay at home orders, for each state. An emphasis was placed on scientific representation of the case numbers, therefore, population doublings (PDs) were calculated on both the log and normal scale (Figure 2C). To highlight and track COVID-19 flattening, a rolling slope was calculated (Figure 2D), showing the slope of the cumulative case regression over all 1-week intervals. Reductions of the rolling slope indicate a flattening of the curve, whereas increases of the slope indicate an acceleration in reported case number. Notably for Los Angeles County a 2-week delay in rolling slope reduction is observed between infections and deaths following issuance of the California Safer at Home order.

This tool can be used to visualize how COVID-19 has spread throughout communities and how state-wide efforts to reduce transmission have affected the curve. Our rolling slope function is an easily digestible method of evaluating changes in direction and rate of confirmed COVID-19 infections and deaths over time. Our goal is to inform people about the spread of this disease and allow them to make more informed decisions after understanding the data. Moreover, the downable nature of the data in our application can facilitate further assessments of trends over time. Assuming that COVID-19 surveillance will continue in earnest throughout the Fall and Winter of 2020, and into the Spring of 2021, we speculate that visualizations such as the rolling slope may help identify resurgence of infections within counties and states

## Availability

The tool can be accessed online (http://usacovid19tracker.com) and the underlying data for county and state level can be directly downloaded through the application.

## Data Availability

Data available through webpage

http://usacovid19tracker.com

## Notes

### Competing Interest Statement

The authors have declared no competing interest.

